# Multi-component Stroke Intervention and Long-term Biofunctional Outcomes: A Secondary Analyses of the SINEMA Trial

**DOI:** 10.64898/2026.01.28.26345092

**Authors:** Bolu Yang, Xiaoyan Yan, Zian Zheng, Fei Wu, Xiong Ding, Xingxing Chen, Brian Oldenburg, Haizhu Song, Yuxuan Zhou, Hanting Zhang, Bifeng Yuan, Lijing L. Yan, Enying Gong

## Abstract

**Background:** The one-year SINEMA trial demonstrated improved blood pressure (BP) control and reduced mortality up to 72 months after the intervention. This article aims to assess between-arm differences in mean annual cumulative BP and to explore whether the associations between cumulative BP and biofunctional outcomes differed by trial arm.

**Methods:** Post-hoc secondary analysis of the SINEMA cluster-randomized trial, which recruited 1299 adults with stroke from 50 rural villages in Hebei, China, between 2017 and 2018. The 12-month intervention was followed by observational assessments at 72 and 84 months post-baseline.

BP was measured during each face-to-face follow-up, assessed by blinded assessors at baseline, 12, 72, and 84 months. Mean annual cumulative systolic BP (SBP), diastolic BP (DBP), mean arterial pressure (MAP), and pulse pressure (PP) were calculated. Biofunctional outcomes included health-related quality of life, modified Rankin Scale, activities of daily living, physical function, and cognition function.

**Results:** Among 897 participants (mean age 62.7 years; 40.8% female) with complete data across all assessment, the intervention arm demonstrated significantly lower mean annual cumulative SBP (−2.2 mm Hg; 95% CI, −3.9 to −0.6), DBP (−1.6 mm Hg; 95% CI, −2.4 to −0.7), and MAP (−1.8 mm Hg; 95% CI, −2.8 to −0.8), not PP, compared with usual care. Significant associations between cumulative BP and biofunctional outcomes were observed in the control arm while not in the intervention arm. Interaction effects between trial arm and cumulative BP were significant for multiple outcomes, most prominently for cumulative SBP.

**Conclusions:** The one-year SINEMA intervention was associated with lower cumulative BP burden over 72-84 months but did not improve overall biofunctional outcomes. Secondary analyses revealed that the association between cumulative BP burden and biofunctional decline differed by intervention arm, suggesting cumulative BP exposure may be an important long-term risk indicator and the intervention may modify BP-outcome relationships through mechanisms requiring further investigation.

## Introduction

Stroke remains a leading cause of long-term disability worldwide, with biofunctional decline encompassing both physical disability and cognitive impairment.^1,2^ Global estimates suggest that approximately 75% of patients with stroke experience some degree of disability, with 15–30% suffering severe impairment.^3^ In China, a large cross-sectional study involving 24,055 patients with first-ever ischemic stroke reported that 78.7% of patients developed post-stroke cognitive impairment.^4^ This decline in physical and cognitive functioning markedly limits the ability to perform daily activities, and the burden becomes more pronounced with aging.^5,6^ Over 20% of patients with stroke are estimated to have limitations in basic activities of daily living (ADLs),^7^ while over 30% experience restrictions in instrumental activities of daily living (IADLs),^8^ with these rates increasing progressively over time.^9^

While emerging evidence suggests associations between elevated blood pressure (BP) and poorer post-stroke biofunctional outcomes including disability measured by modified Rankin Scale (mRS)^10^ and cognitive functions^11,12^, findings remain inconsistent across studies and biofunctional domains. Evidence is limited for the association between BP and other critical domains including activities of daily living (ADL), physical function, and health-related quality of life. Additionally, while studies have demonstrated that biofunctional decline is more strongly associated with poor long-term BP control,^13,14^ a key limitation of existing research is that most studies have assessed BP at single time points, ^15–18^ which may not fully capture the long-term burden of BP exposure on biofunctional outcomes. Cumulative BP, as a measure that integrates BP levels over extended periods, has emerged as a potentially important metric for associations with cardiovascular disease risk. ^19^ However, whether cumulative BP differentially affects various post-stroke biofunctional domains remains unclear, and evidence linking cumulative BP to comprehensive biofunctional outcomes is largely lacking.

The SINEMA (<u>s</u>ystem-integrated and tech<u>n</u>ology-<u>e</u>nabled <u>m</u>odel of c<u>a</u>re) trial was designed to improve BP control among patients with stroke in resource-limited rural settings in China.^20^ The trial showed significant improvements in BP control during the 12-month intervention period through a multifaceted strategy combining digital health tools and primary care support.^21,22^ To address the lack of long-term evidence of the cumulative BP and biofunctional outcomes, the trial incorporated follow-up assessments at 72 and 84 months post-baseline. In this post-hoc secondary analysis, we aimed to (1) quantify and compare mean annual cumulative BP between trial arms through 84 months, and (2) examine whether the associations between cumulative BP and biofunctional outcomes differed by intervention arm.

## Methods

### Data Availability

Deidentified individual data can be made available upon written, detailed request directed to the corresponding author. Requests will be reviewed on the basis of scientific merit, ethical review, legal issues, and regulatory requirements.

### Study Design and Participants

This study is a post-hoc secondary analysis of the SINEMA trial. The original trial was a 12-month cluster-randomized controlled trial, followed by observational follow-up assessments at 72- and 84-months post-baseline. The detailed trial design ^20^, digital health intervention components^21^, and primary within-trial^22^ and post-trial outcomes^23–25^ have been described previously. Eligible participants were adults with a confirmed history of stroke who were in a clinically stable condition. Between June 23 and July 21, 2017, a total of 1,299 patients with stroke were enrolled and followed for 12 months during the intervention period. Following completion of the trial’s intervention phase, long-term follow-up assessments were conducted in 72- and 84-months post-baseline to evaluate the sustained effects of the intervention (***eSupplement 1***).

In brief, the SINEMA trial was a cluster-randomized controlled trial conducted in rural Hebei Province, China. A total of 50 villages from five townships were randomized in a 1:1 ratio, stratified by township, to either the intervention arm or the usual care control arm. In the intervention arm, village doctors underwent training, were equipped with the SINEMA app, and performed monthly follow-up visits for patients with stroke with support from township physicians and financial incentives. Patients with stroke received these visits either at village clinics or at home and received daily voice messages containing health education on medication adherence and physical activities. The intervention ran from June 23, 2017, to July 31, 2018. After support ended, village doctors could voluntarily continue using the SINEMA app free of charge and provide services without incentives. In the control arm, participants received usual care and had to seek care proactively; patients with hypertension and/or type 1 or 2 diabetes were eligible for quarterly follow-ups and general health education under the national public health program.^26^

### Assessment of Blood Pressure and Mean Annual Cumulative BP Calculation

The predefined primary outcome of the SINEMA trial was SBP. In this post-hoc secondary analysis, we additionally evaluated diastolic BP, mean arterial pressure (MAP), and pulse pressure (PP), with MAP and PP derived using standard formulas: PP = SBP − DBP; MAP = DBP + 1/3× PP. BP was measured on the right upper arm with participants seated and resting for at least 5 minutes, using a validated electronic sphygmomanometer (*Omron HEM-7052*). Two measurements were obtained, and if the difference between the two SBP readings exceeded 10 mmHg, a third measurement was taken. The mean of the two closest readings was used in all analyses (***eSupplement 1***).

Cumulative BP exposure was approximated by the area under the BP–time curve (AUC) via the trapezoidal rule and then divided by the corresponding follow-up duration (6 years for the 72-month analyses; 7 years for the 84-month analyses) to obtain the mean annual cumulative BP (units: mmHg), reflecting average long-term BP burden. Because intermediate years lacked planned measurements, we used only the available time points without imputation. The calculation for the 84-month analysis is illustrated below:

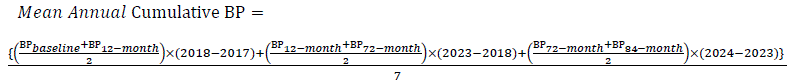

### Measurement on biofunctional outcomes

Biofunctional outcomes encompassed overall function status (quality of life, ADL, IADL, and disability measured by the modified Rankin Scale [mRS]), physical function (upper limb grip strength and lower limb performance), and cognitive function.

Quality of life was assessed using the EQ-5D-5L, scored with the Chinese value set.^27,28^ ADL (range 0–6) and IADL (range 0–8) were assessed using a modified Katz and Lawton scale widely applied in Chinese aging studies, with higher scores indicating greater dependence.^29,30^ Disability was assessed with the mRS (range 0–6), with higher scores indicating greater disability.^31^ For physical function, upper limb function was assessed using grip strength, measured with a WL-1000 dynamometer. Each participant performed two trials with each hand, and the maximum value was recorded. Participants unable to perform the test were assigned a value of 0 kg. Lower limb function was evaluated using the Timed Up and Go (TUG) test. Cognitive function was evaluated with the brief Community Screening Instrument for Dementia (CSI-D, score range 0–7).^32^ Data on patients’ self-reported information was collected through face-to-face interviews and entered directly into an electronic data capture system (*Qualtrics, Provo, Utah*). All assessments were performed by trained data collectors blinded to arm allocation, with detailed protocols provided in ***eSupplement 1***.

### Statistical Analysis

Baseline characteristics were summarized by arm, using mean (standard deviation, SD) for continuous variables and frequency (percentage) for categorical variables. Analyses were conducted in the intention-to-treat at individual level, accounting for clusters and stratified design per the pre-published statistical analysis plan (SAP).^33^ Between-arm differences on mean annual cumulative BP, BP and biofunctional outcomes at 72 and 84 month follow-up visits were estimated using mixed-effects models, incorporating a random intercept for cluster (village) and fixed effect for townships, baseline outcomes, age, baseline DBP and sex.^33^ For binary outcomes, we employed generalized estimating equations (GEE) with a Poisson distribution and log link, specifying robust standard errors and an independent correlation structure. Results are reported as risk ratios (RR).^34^

To assess whether the intervention modified the association between cumulative BP and biofunctional outcomes, we included an interaction term (cumulative BP × intervention arm) in the model. Biofunctional outcomes from 84-month follow-up were used. Quality of life was analyzed as a continuous variable. To better capture gradations of function, ADL and IADL were analyzed as count scores, reflecting the number of biofunctional limitations.^6^ Disability was dichotomized using the mRS as ≤3 versus >3.^35,36^ TUG test dichotomized at ≥14 seconds or inability to complete. Reduced grip strength was defined using sex-specific median values derived from our study population (<28 kg for men; <16 kg for women), consistent with prior literature.^37^ Cognitive function was dichotomized at a cutoff score of ≤6, which has been validated in the Chinese population with a sensitivity of 81.3% and specificity of 95.1%, to indicate cognitive impairment.^32^ For ADL and IADL, residuals did not meet the normality assumption; therefore, bootstrapping with 1,000 replications clustered at the village level was applied, consistent with the SAP. ^33^ For grip strength, ADL, IADL, and cognitive function, baseline values were not available and therefore were not included as covariates in the models.

Several sensitivity analyses were conducted. First, we used mean annual cumulative BP up to 84 months and tested its interaction with intervention arm on 84-month biofunctional outcomes. Second, we excluded participants with extreme values, defined as those below the 25th percentile minus two times the interquartile range or above the 75th percentile plus two times the interquartile range. Third, multiple imputations using chained equations (MICE) were applied to address missing data on SBP, DBP, and biofunctional outcomes at the 72- and 84-month assessments, restricted to participants alive at each assessment because biofunctional outcomes were only measurable among survivors; 20 imputed datasets were generated. Finally, to address potential nonlinearity in BP trajectories and sensitivity to the linear interpolation assumption in the primary analysis, we used restricted cubic spline (RCS) mixed-effects models were used to predict missing SBP values and construct continuous individual SBP trajectories. Mean annual cumulative SBP was then recalculated using trapezoidal integration of the predicted values.

## Results

A total of 1,299 participants from 50 villages were enrolled at baseline and 1,226 participants were followed at the end of the trial at 12 months, with 30 deaths and 43 individuals lost to follow-up (3.4%). By 72 months post-baseline assessment, 227 participants had died and 44 (4.2%) were lost to follow-up. At 84 months, 912 participants completed follow-up, with 65 participants died in one year and 81 individuals (8.1%) lost to follow-up. 897 participants with complete data at both 72 and 84 months were included in the analysis (***Figure 1***). Baseline characteristics of 897 participants are summarized in ***Table 1***. The mean (standard deviation [SD]) baseline age was 65 (8) years, and 46% of participants were female. The mean baseline SBP and DBP were 146 (21) mmHg and 79 (12) mmHg, respectively. Baseline DBP was slightly higher in the intervention arm (mean [SD], 79.8 [11.3] mm Hg) than in the control arm (78.2 [11.8] mm Hg) and was adjusted for in all multivariable analyses. Other baseline characteristics were balanced between arms. Other baseline demographic and biofunctional characteristics were balanced between the two arms.

**Figure 1.**
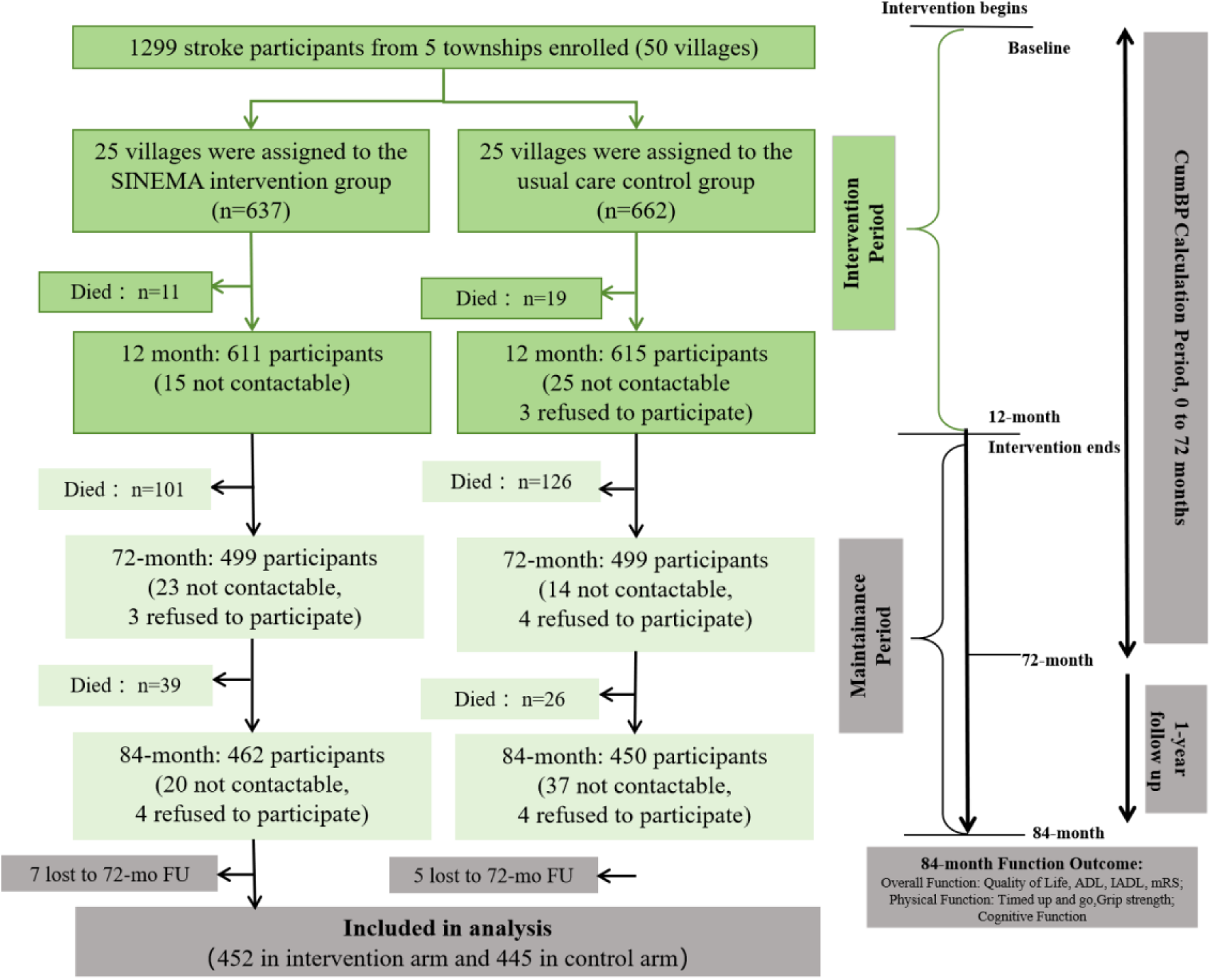
Flow diagram of participants through enrollment, randomization, and follow-up.

**Table 1.**
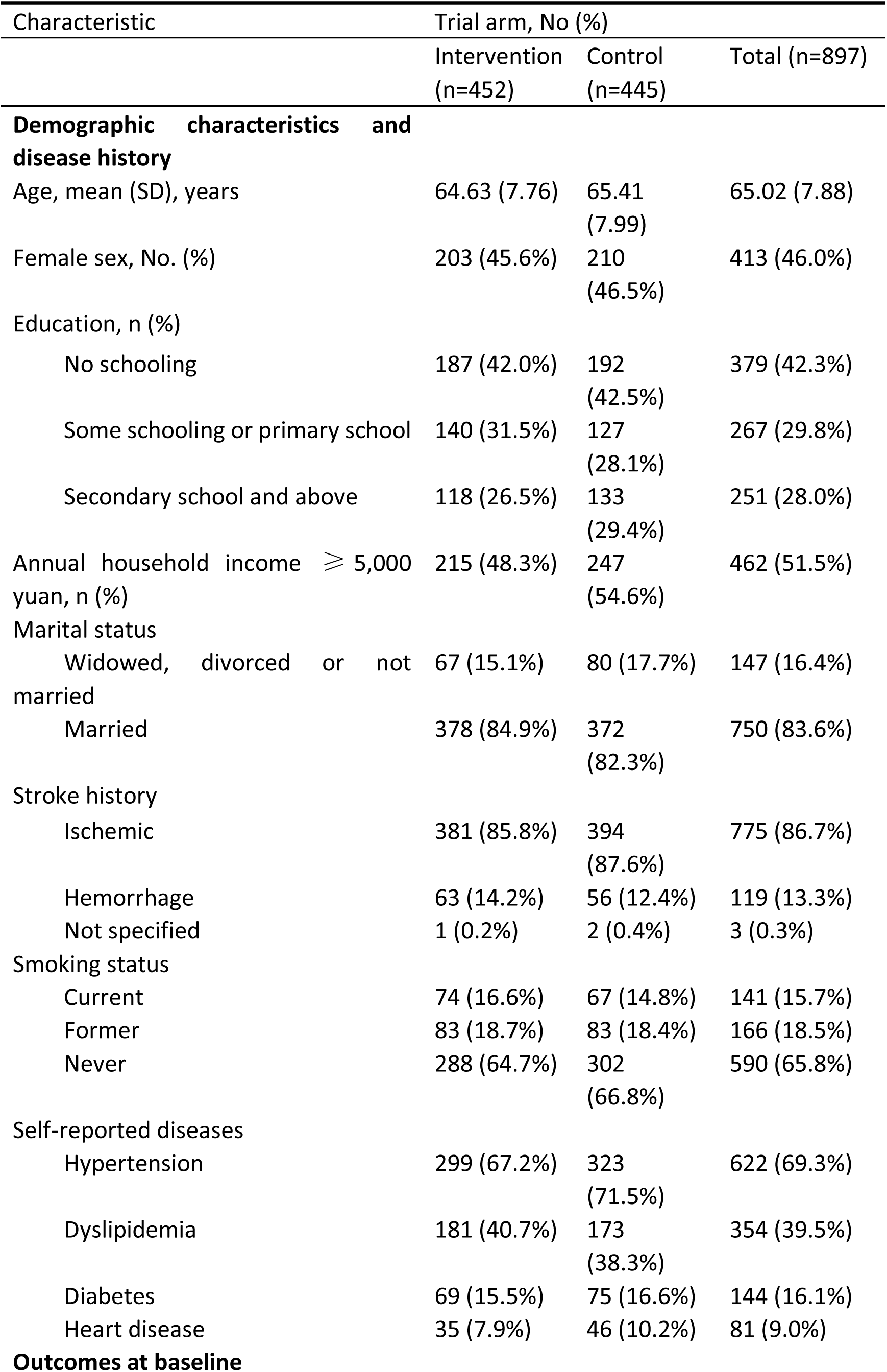
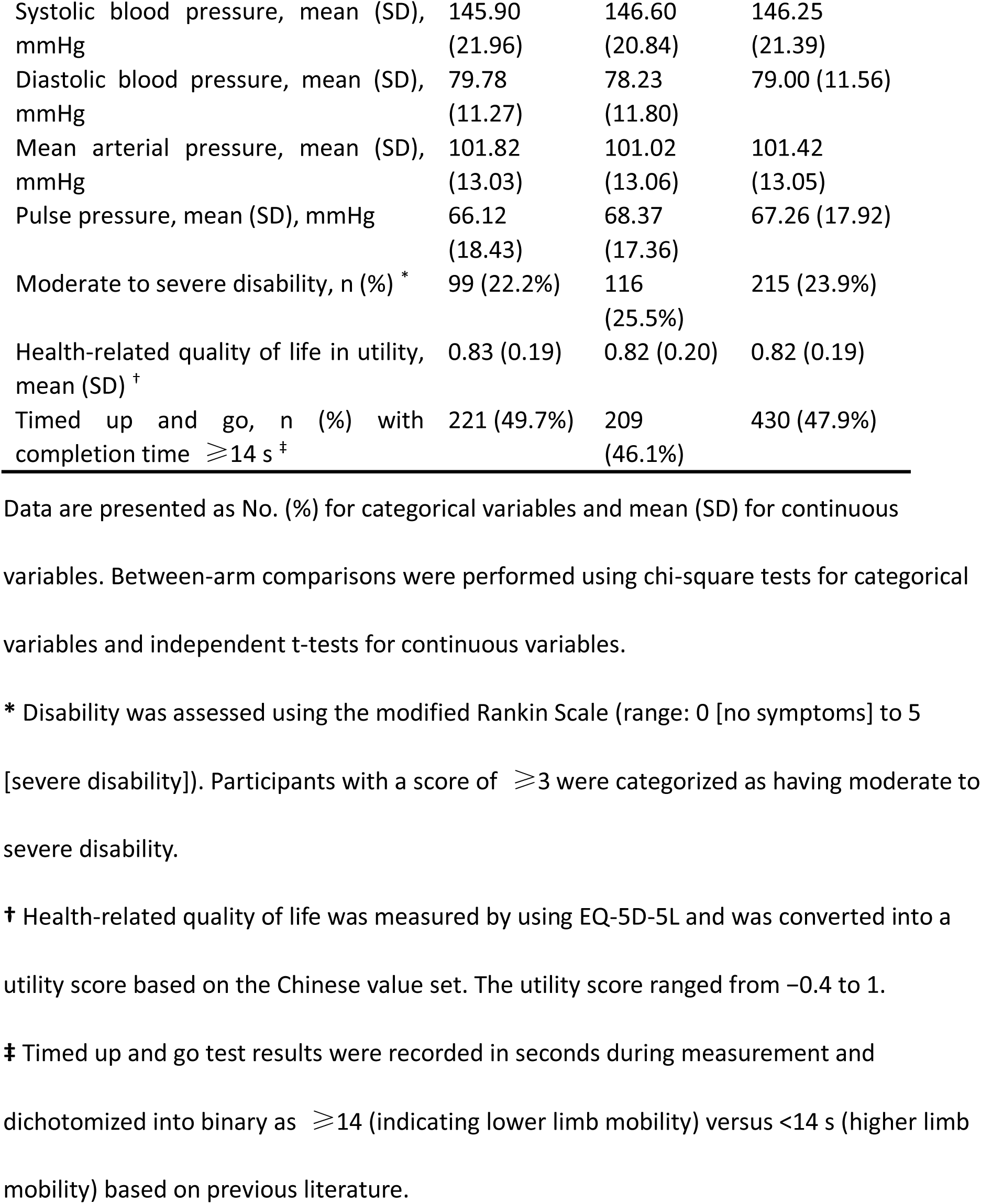
Baseline Characteristics of Study Participants.

| Characteristic | Trial arm, No (%) |  |  |
| --- | --- | --- | --- |
|  | Intervention<br>(n=452) | Control<br>(n=445) | Total (n=897) |
| **Demographic characteristics and disease history** |  |  |  |
| Age, mean (SD), years | 64.63 (7.76) | 65.41<br>(7.99) | 65.02 (7.88) |
| Female sex, No. (%) | 203 (45.6%) | 210<br>(46.5%) | 413 (46.0%) |
| Education, n (%) |  |  |  |
| No schooling | 187 (42.0%) | 192<br>(42.5%) | 379 (42.3%) |
| Some schooling or primary school | 140 (31.5%) | 127<br>(28.1%) | 267 (29.8%) |
| Secondary school and above | 118 (26.5%) | 133<br>(29.4%) | 251 (28.0%) |
| Annual household income ≥ 5,000 yuan, n (%) | 215 (48.3%) | 247<br>(54.6%) | 462 (51.5%) |
| Marital status |  |  |  |
| Widowed, divorced or not married | 67 (15.1%) | 80 (17.7%) | 147 (16.4%) |
| Married | 378 (84.9%) | 372<br>(82.3%) | 750 (83.6%) |
| Stroke history |  |  |  |
| Ischemic | 381 (85.8%) | 394<br>(87.6%) | 775 (86.7%) |
| Hemorrhage | 63 (14.2%) | 56 (12.4%) | 119 (13.3%) |
| Not specified | 1 (0.2%) | 2 (0.4%) | 3 (0.3%) |
| Smoking status |  |  |  |
| Current | 74 (16.6%) | 67 (14.8%) | 141 (15.7%) |
| Former | 83 (18.7%) | 83 (18.4%) | 166 (18.5%) |
| Never | 288 (64.7%) | 302<br>(66.8%) | 590 (65.8%) |
| Self-reported diseases |  |  |  |
| Hypertension | 299 (67.2%) | 323<br>(71.5%) | 622 (69.3%) |
| Dyslipidemia | 181 (40.7%) | 173<br>(38.3%) | 354 (39.5%) |
| Diabetes | 69 (15.5%) | 75 (16.6%) | 144 (16.1%) |
| Heart disease | 35 (7.9%) | 46 (10.2%) | 81 (9.0%) |
| **Outcomes at baseline** |  |  |  |
| Systolic blood pressure, mean (SD), mmHg | 145.90 (21.96) | 146.60 (20.84) | 146.25 (21.39) |
| Diastolic blood pressure, mean (SD), mmHg | 79.78 (11.27) | 78.23 (11.80) | 79.00 (11.56) |
| Mean arterial pressure, mean (SD), mmHg | 101.82 (13.03) | 101.02 (13.06) | 101.42 (13.05) |
| Pulse pressure, mean (SD), mmHg | 66.12 (18.43) | 68.37 (17.36) | 67.26 (17.92) |
| Moderate to severe disability, n (%) * | 99 (22.2%) | 116 (25.5%) | 215 (23.9%) |
| Health-related quality of life in utility, mean (SD) † | 0.83 (0.19) | 0.82 (0.20) | 0.82 (0.19) |
| Timed up and go, n (%) with completion time $\geq 14$ s ‡ | 221 (49.7%) | 209 (46.1%) | 430 (47.9%) |

Data are presented as No. (%) for categorical variables and mean (SD) for continuous variables. Between-arm comparisons were performed using chi-square tests for categorical variables and independent t-tests for continuous variables.
\* Disability was assessed using the modified Rankin Scale (range: 0 [no symptoms] to 5 [severe disability]). Participants with a score of $\geq 3$ were categorized as having moderate to severe disability.
† Health-related quality of life was measured by using EQ-5D-5L and was converted into a utility score based on the Chinese value set. The utility score ranged from -0.4 to 1.
‡ Timed up and go test results were recorded in seconds during measurement and dichotomized into binary as $\geq 14$ (indicating lower limb mobility) versus $<14$ s (higher limb mobility) based on previous literature.

As shown in ***Figure 2***, the intervention arm had consistently lower mean annual cumulative BP compared with the control arm at both follow-up time points. At 72 months, the intervention arm had significantly lower cumulative SBP (−2.21 mm Hg; 95% CI, −3.86 to −0.55 mm Hg), DBP (−1.56 mm Hg; 95% CI, −2.37 to −0.74 mm Hg), and MAP (−1.76 mm Hg; 95% CI, −2.77 to −0.75 mm Hg). Similar patterns were observed at 84 months for SBP (−1.95 mm Hg; 95% CI, −3.61 to −0.28 mm Hg), DBP (−2.33 mm Hg; 95% CI, −3.42 to −1.23 mm Hg), and MAP (−2.19 mm Hg; 95% CI, −3.34 to −1.05 mm Hg). No significant between-arm differences were observed for cumulative PP at either time point (72 months: −0.49 mm Hg; 95% CI, −1.72 to 0.75 mm Hg; 84 months: 0.39 mm Hg; 95% CI, −1.11 to 1.88 mm Hg).

**Figure 2.**
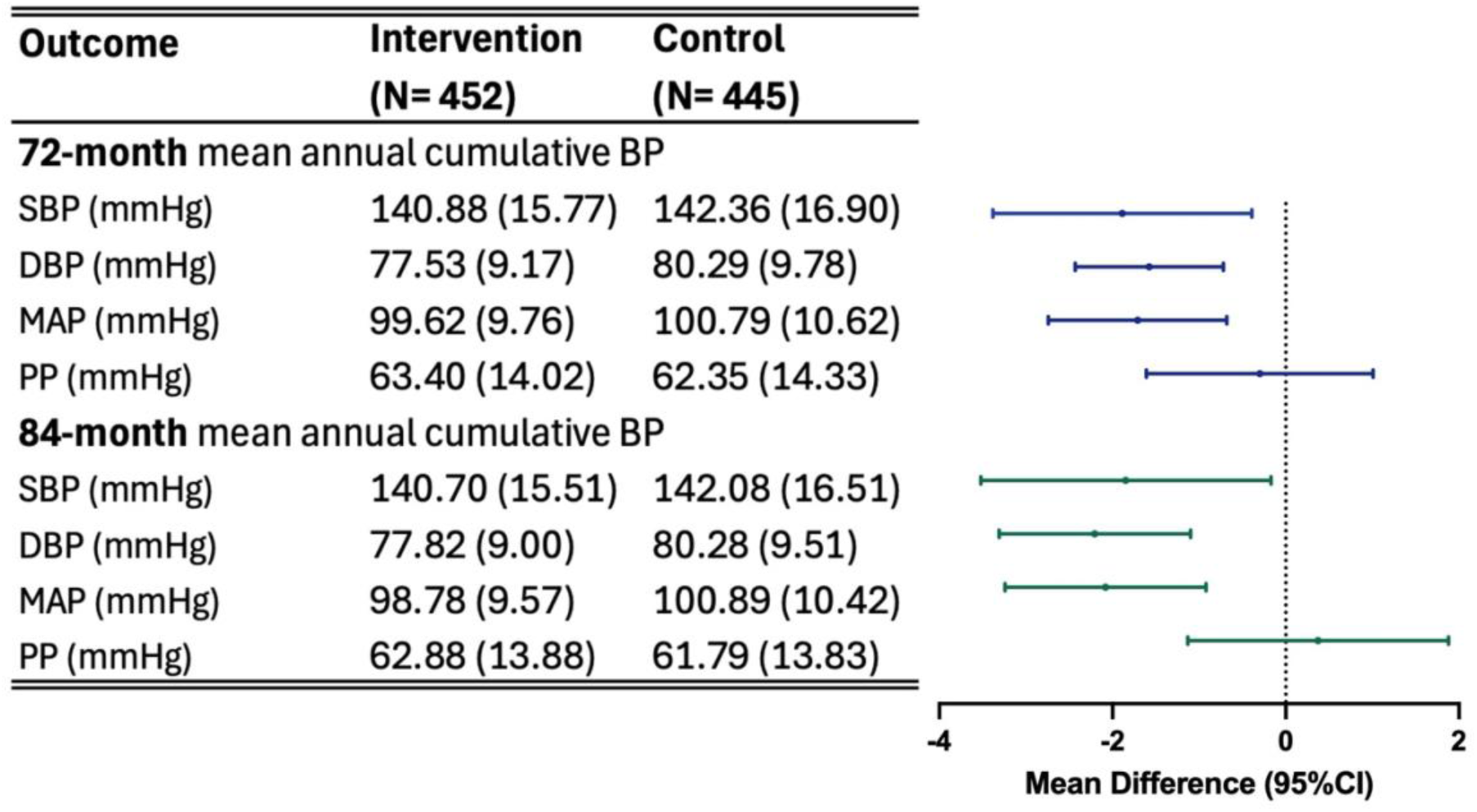
Between-Arm Differences in Mean Annual Cumulative Blood Pressure at 72 and 84 Months. Data are presented as mean (standard deviation, SD) and mean difference (95% confidence interval, CI) between the two arms from baseline to 72 months. BP, blood pressure; MAP, mean arterial pressure; PP, pulse pressure. Mean difference is estimated using mixed-effects models with a random intercept for cluster (village) to account for the cluster-randomized design, and fixed effects for township (to account for stratified randomization), baseline outcome values, age, baseline DBP, and sex

In the overall cohort, higher mean annual cumulative BP was associated with poorer biofunctional outcomes, with the strength and consistency of associations varying by BP component and outcome domain (***eTable 1 in Supplement 2***). Cumulative DBP demonstrated the most consistent associations across all biofunctional outcomes except grip strength, whereas cumulative SBP showed more selective effects, with significant associations limited to quality of life, mRS and IADL. Neither cumulative SBP nor DBP was significantly associated with grip strength. Cumulative PP showed no significant associations with any biofunctional outcome. Stratified analyses revealed substantial differences in these associations by trial arm. In the control arm, significant associations between higher cumulative BP and poorer biofunctional outcomes were consistently observed across multiple domains. In contrast, these associations were largely attenuated in the intervention arm (***Figures 3-4***). Interaction analyses confirmed that the associations between cumulative BP and biofunctional outcomes were modified by intervention arm, with patterns differing by BP component (***Figures 3-4***). Cumulative SBP showed significant interactions for 7 outcomes (all P ≤ 0.05), with the strongest observed for quality of life (P = 0.003) and the weakest for grip strength (P = 0.048). Cumulative PP demonstrated a similar pattern across six outcomes. In contrast, cumulative DBP showed no significant interactions, while cumulative MAP showed a significant interaction only for quality of life (P = 0.007). These differential associations were observed despite similar absolute biofunctional outcome levels between arms at 72 and 84 months (***eTable 2 in Supplement 2***).

**Figure 3.**
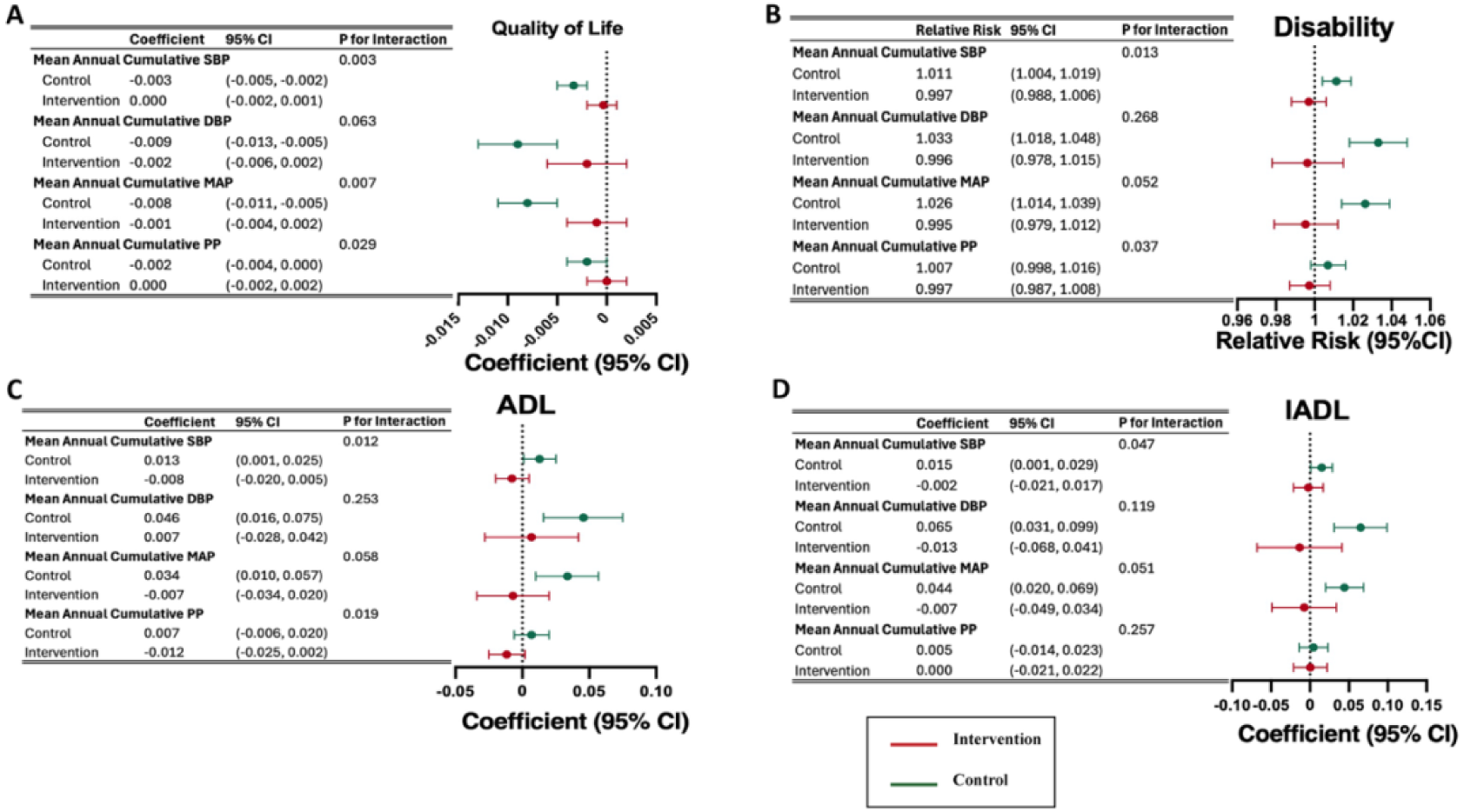
Associations Between Cumulative Blood Pressure and Overall Functional Outcomes by Arm. BP, blood pressure; DBP, diastolic blood pressure; IADL, instrumental activities of daily living; MAP, mean arterial pressure; mRS, modified Rankin Scale; PP, pulse pressure; SBP, systolic blood pressure. *. Mean annual cumulative blood pressure was calculated up to 72 months, while functional outcomes were assessed at 84 months. Estimates were obtained from mixed-effects models accounting for village clustering and adjusted for township stratification, baseline outcome (where available), age, baseline diastolic blood pressure, and sex. For ADL, IADL—measured only at the 84-month follow-ups—models were not adjusted for baseline ADL and IADL. †. Figure 3A: Health-related quality of life, assessed with EQ-5D-5L and converted to utility scores using the Chinese value set. ‡. Figure 3B: Disability, assessed with the modified Rankin Scale (mRS). Participants with scores >3 were classified as having moderate to severe disability. §. Figure 3C: ADL was analyzed as a continuous variable (range: 0–6). A score of 0 indicates no limitations; 6 indicates complete dependence in all activities. ||. Figure 3D: IADL was analyzed as a continuous variable (range: 0–8). A score of 0 indicates no limitations; 8 indicates complete dependence in all activities.

**Figure 4.**
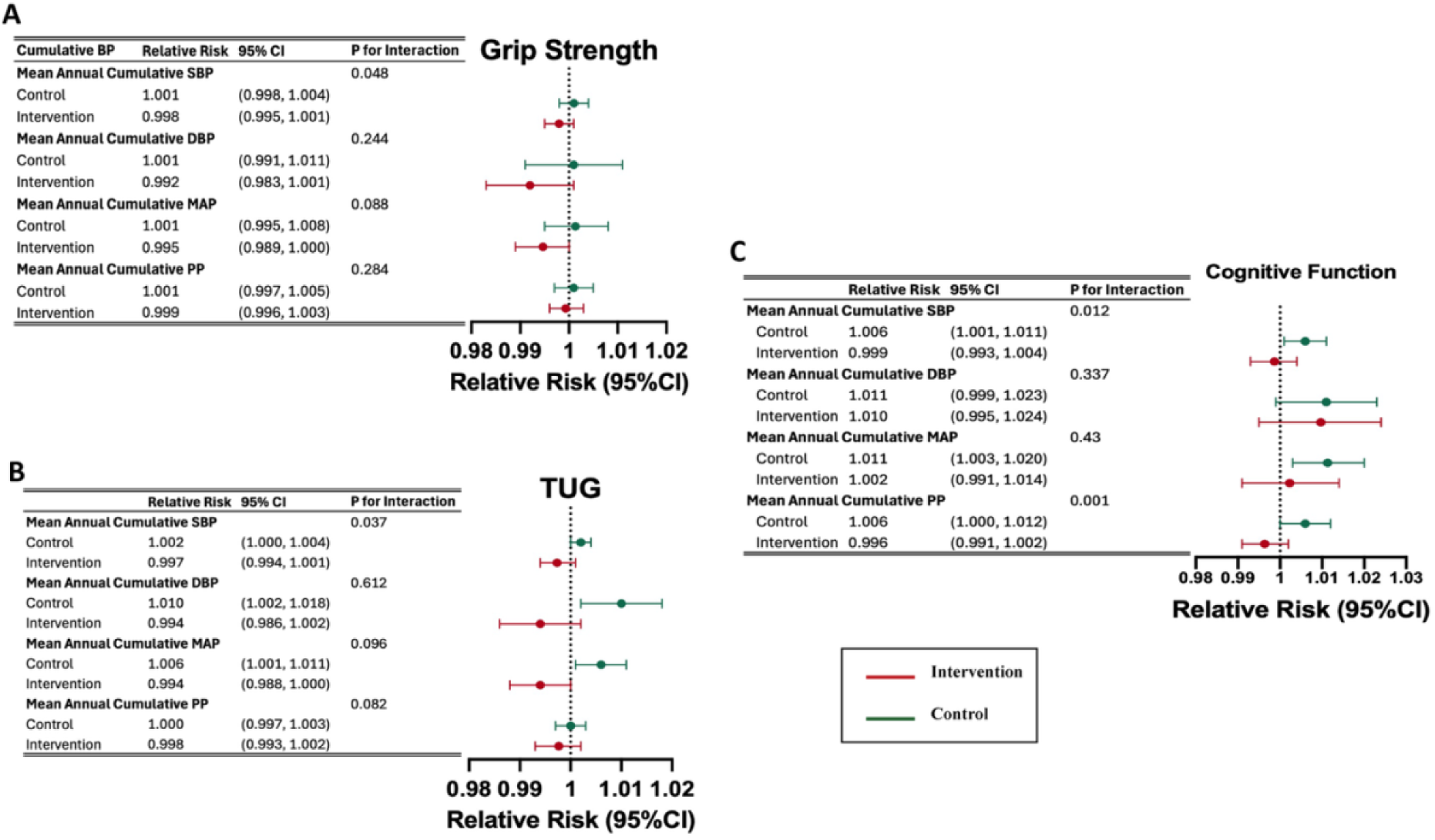
Associations Between Cumulative Blood Pressure and Physical and Cognitive Function Outcomes by Arm. BP indicates blood pressure; CSI-D, Community Screening Instrument for Dementia; DBP, diastolic blood pressure; MAP, mean arterial pressure; PP, pulse pressure; SBP, systolic blood pressure; TUG, Timed Up and Go test. *. Mean annual cumulative blood pressure was calculated up to 72 months, while functional outcomes were assessed at 84 months. Estimates were obtained from mixed-effects models accounting for village clustering and adjusted for township stratification, baseline outcome (where available), age, baseline diastolic blood pressure, and sex. For grip strength, cognitive function—measured only at the 84-month follow-ups—models were not adjusted for baseline grip strength and cognitive function. †. Figure 4A: Grip strength, analyzed as a categorical variable using sex-specific cutoffs (<28 kg for men, <16 kg for women). Participants unable to perform the test were assigned a value of 0 kg. ‡. Figure 4B: Lower limb strength, assessed with the TUG test. Results (in seconds) were dichotomized as ≥14 (impaired lower limb mobility) vs <14 (normal mobility). §. Figure 4C: Cognitive function, assessed using the Community Screening Instrument for Dementia (CSI-D). Impairment was defined as a total score ≤6.

### Sensitivity Analysis

Results were generally robust across sensitivity checks. When substituting 84-month mean annual cumulative BP for the 84-month function outcomes in the interaction models, estimates were stable overall (***eFigures 1 and 2 in Supplement 2***). After excluding outliers, the intervention arm continued to show significant differences in mean annual cumulative BP at both the 72- and 84-month follow-ups (***eTable 3 in Supplement 2***), and interaction patterns remained consistent (***eFigures 3 and 4 in Supplement 2***). Multiple imputation analyses for missing BP and biofunctional outcome data yielded similar findings (***eTable 4 and eFigures 5-7 in Supplement 2***). Analyses using restricted cubic spline models to predict BP values at unmeasured time points showed consistent interactions between intervention arm and cumulative SBP for quality of life; interactions for other outcomes were directionally similar but did not reach statistical significance (***eTable 5 and eFigures 7 and 8 in Supplement 2***).

## Discussion

Building on prior evidence of sustained BP control after the SINEMA intervention,^24^ this post-hoc analysis demonstrates lower cumulative SBP, DBP, and MAP in the SINEMA intervention arm compared with control arm over seven years of follow-up. Higher cumulative BP was consistently associated with poorer biofunctional outcomes in the usual care l arm, whereas these associations were largely attenuated in the intervention arm, with significant interactions between cumulative BP and intervention observed for multiple outcomes. These findings suggest that the SINEMA intervention may modify the relationship between long-term BP burden and biofunctional decline among stroke survivors.

Cumulative BP integrates both the intensity and duration of exposure and may better reflect long-term vascular burden than single time-point measurements.^38^ However, few secondary prevention trials have evaluated intervention effects using cumulative BP metrics. For instance, the SSaSS trial assessed the effectiveness of salt substitute on various cumulative BP during the active intervention period, illustrating the observed differences in cumulative SBP, MAP, PP but not DBP.^39^ By extending follow-up to 72 and 84 months, our findings add to this evidence by showing that the SINEMA intervention could not only improve BP control within the trial period, but also were associated with sustained reductions in cumulative SBP, DBP, MAP beyond the intervention period. The absence of a significant differences in cumulative PP aligns with emerging evidence from cohort study suggesting limited predictive utility of cumulative PP for cardiovascular outcomes in Chinese populations.^40^

A key finding of the analysis is that cumulative BP was strongly associated with biofunctional decline across multiple domains, including quality of life, disability, activity of daily living, cognitive function, physical function in the usual care arm, but not in the intervention arm. The observed association for usual care arm extend prior work that predominantly focused on BP associations with cognitive function or basic disability measures.^12,13^ Interaction analyses supported effect modification by intervention status, particularly for cumulative systolic blood pressure. These findings suggest that the SINEMA intervention may have mitigated the adverse biofunctional consequences typically associated with prolonged BP exposure among stroke survivors.

The observed interaction effect but similar absolute level of biofunctional outcomes may be explained by the magnitude of the BP reduction during the post-trial period. The observed reduction in BP for was smaller than 10-15 mmHg reductions typically required to produce detectable biofunctional or cognitive benefits in prior trials.^41,42^ Such observed pattern from post-trial analysis indicates that the sustained effect of the SINEMA intervention on cumulative BP may not be sufficiently intensive to produce detectable population-level improvements in function, however, it altered the relationship between BP burden and biofunctional decline. Such effect modification is clinically meaningful, as it suggests that the SINEMA models may confer resilience against vascular risk–related biofunctional deterioration, even in the absence of large average reductions in disability or impairment.

Interestingly, the interaction patterns differed substantially by BP components. Cumulative SBP and PP showed significant interactions with intervention arms across all or 6 biofunctional domains, whereas no significant interactions between DBP and intervention on any of the biofunctional outcomes and cumulative MAP showed significant interaction with the intervention only for quality of life. These findings are exploratory in post-hoc analysis but suggest that long-term SBP may be particularly relevant to biofunctional outcomes after stroke and may be more amenable to modification through SINEMA intervention. If confirmed in other studies, these results could help refine BP targets in secondary stroke prevention by emphasizing cumulative SBP rather than isolated measurement.

This study has several important public health and clinical implications. First, our findings suggest that cumulative BP burden rather than single time-point measurements may be a more informative target and monitoring metric for long-term stroke management, complementing the most recent clinical guidelines for stroke secondary prevention.^43^ Second, the differential associations between cumulative BP and biofunctional decline in the intervention and control arms, even without detectable overall biofunctional benefit, raise the possibility that the SINEMA model may protect against BP-related biofunctional decline through mechanisms beyond BP lowering alone, such as improved self-management skills, health literacy and care continuity. However, given the exploratory nature of these interaction analyses, these findings require validation in future studies. Third, the association between cumulative BP exposure and long-term biofunctional outcomes emerged only through extended follow-up beyond the active intervention period, highlighting the critical value of long-term monitoring in illustrating the full impact of secondary stroke prevention strategies.

## Limitations

Our study has several limitations. First, as a post-hoc exploratory secondary analysis, this study was not designed with prespecified hypotheses regarding cumulative BP and biofunctional outcomes and are subject to multiple testing. Accordingly, these findings should be interpreted in caution and causal inference is limited by the observational nature of the post-trial follow-up. Second, survivor bias is inevitable as biofunctional outcomes were assessed only among participants alive at follow-up, which may underestimate the observed associations. Third, several biofunctional measurements, such as grip strength, ADL/IADL, and cognitive function, were assessed only at post-trial follow-up, precluding baseline adjustment for these outcomes, although randomization ensured balance in baseline characteristics and imbalanced factors were adjusted in the analyses. Finally, detailed data on post-trial intervention adherence and more frequent BP monitoring were unavailable, limiting assessment of potential mediating pathways; therefore some of the findings warrant confirmation in future studies.

## Conclusion

In this post-hoc secondary analysis of the SINEMA trial, a digital health-enabled, primary-care based stroke intervention was associated with lower long-term cumulative BP over 84 months of follow-up and with attenuation of the association between cumulative BP and poorer biofunctional outcomes across multiple domains. These findings highlight cumulative BP as a potentially important long-term risk indicator and suggest that the SINEMA model may contribute to biofunctional preservation among stroke survivors. Confirmation in prospectively designed studies is warranted with further evaluation of the generalizability and scalability of this model in other settings.

## Acknowledgments

We would like to thank the independent International Steering Committee Chair (Yangfeng Wu) and members (Eric Peterson and Craig Anderson) and advisory arm members (Alba AmayaBurns, Allan Burns, Ninghua Wang, Xie Bin, Jesse Hao, Jixiang Ma, Jixin Sun, Jianxin Zhang, Jianmin Yao, Jinmei Liu, Qian Long, and Cheng Sun) who have provided great advice in designing and implementing the study. We also thank the Xingtai City Center for Disease Prevention and Control, Nanhe County Center for Disease Prevention and Control, Ren County Center for Disease Prevention and Control, all staff members from township health care centers and village clinics, and all patients who participated in or supported the project.

## Supplemental Material

Trial and Follow-up Protocol

Tables S1–S5

Figure S1-S8

CONSORT guidelines

